# Decision models to improve healthcare-associated *C. difficile* PCR test use in patients with community-acquired pneumonia

**DOI:** 10.1101/2020.08.25.20181925

**Authors:** Nathaniel J. Rhodes, Caroline C. Jozefczyk, W. Justin Moore, Paul R. Yarnold, Karolina Harkabuz, Sarah H. Sutton, Christina Silkaitis, Chao Qi, Richard G. Wunderink, Teresa R. Zembower

## Abstract

Adults hospitalized with community-acquired pneumonia (CAP) typically receive antibiotics and thus are at increased risk of developing *Clostridioides difficile* infection (CDI), a disease of significant morbidity. We developed and validated a CAP-specific clinical decision algorithm to facilitate optimal diagnostic stewardship of *C. difficile* polymerase chain reaction (PCR) testing. The study was a single-center retrospective, case-control analysis of hospitalized adult patients empirically treated for CAP between January 1, 2014 and May 29, 2018. A series of predictive models and validity assessments were used to evaluate demographic and post-admission patient-specific risk factors as predictors of CDI case status among patients with CAP. Thirty-two PCR confirmed CDI cases were identified and 232 randomly selected controls were drawn from the total CAP population. After propensity score weighting, hospital-onset (HO) CDI was significantly associated with broad-spectrum Gram-negative antibiotic use (P=0.002) as was subsequent community-onset (CO) CDI (P=0.005). Modified-APACHE II > 8.5 (P=0.003) and broad-spectrum Gram-negative antibiotic use (P=0.002) were associated with healthcare-associated CDI and were robust in multiple validity analyses. Patients with m-APACHE II ≤ 8.5 who received broad-spectrum Gram-negative antibiotics were more likely (odds=1:2) to experience healthcare-associated CDI compared to those who did not receive these broad-spectrum agents (odds=1:125) and compared to those with m-APACHE II > 8.5 irrespective of treatment (odds=5:27). We conclude that broad-spectrum Gram-negative antibiotic use was the common factor in development of CDI in patients with CAP in all settings. Prospective studies are needed to confirm the reproducibility and clinical utility of our model when used for diagnostic test stewardship.

## Introduction

Pneumonia is the leading infectious cause of death and eighth leading cause of death overall among patients in the U.S. (1). Overuse of broad-spectrum antibiotics, which are likely inappropriate for the majority of patients with community acquired pneumonia (CAP), is a major driver of infectious complications including *Clostridioides difficile* associated diarrhea (2). *C. difficile* infection (CDI) is a significant complication of antibiotic treatment that is responsible for high morbidity. CDI most commonly presents as new onset diarrhea among patients with recent antibiotic exposure, but it can rarely present as ileus and toxic megacolon (3). The diagnosis of CDI relies on both clinical and microbiological evidence of disease. Strictly from an analytical perspective, *C. difficile* polymerase chain reaction nucleic acid amplification testing (i.e., PCR testing) on liquid stool specimens is a highly sensitive and specific test [90% and 96%, respectively (4)]; however, clinical false positive tests can occur in patients who are colonized by *C. difficile* (5, 6). We and others have found inconsistent documentation of clinical disease and a high occurrence of alternate etiologies for diarrhea in hospitalized patients (e.g., laxative use, oral contrast, systemic chemotherapy) (7). To address this gap in practice and thus reduce clinical false positive results, consensus guidelines recommend implementing institutional testing criteria or, alternatively, following a multistep laboratory testing algorithm (3). While the use of institutional testing criteria theoretically should improve PCR test utilization, applying one set of criteria to all hospitalized patients may constitute optimal test use. Thus, a need exists for population-specific test criteria in order to steward *C. difficile* PCR testing.

Diagnostic stewardship within hospitals can be achieved through an integrated approach supported by Antimicrobial Stewardship Programs (ASPs), Infection Prevention Programs, and the clinical microbiology laboratory. Once population-specific risk factors are defined, models that support clinical decision making can help narrow the pool of patients who require PCR testing for definitive diagnosis and prioritize testing for those who are high risk. Certain patient populations may carry higher risk than others, particularly those who often receive antibiotics and have frequent need for hospitalization. Inappropriate antibiotic use in patients with CAP (e.g., excessive duration of therapy) can lead to CDI (8-10). Epidemiological studies have documented a lack of judicious prescribing among patients with lower respiratory tract infections including CAP (11, 12). Inappropriate prescribing corresponds with prolonged lengths of stay (13) and greater mortality (14). Importantly, healthcare-associated outbreaks of CDI have been associated with inappropriate antibiotic use in CAP (15). Therefore, improvements in antibiotic prescribing are expected to both significantly improve outcomes in CAP and reduce the risk of CDI development. The present study develops and validates CAP-specific decision models to support optimal diagnostic and antimicrobial stewardship in this population.

## Materials and Methods

### Design

This study was reviewed and approved by the Institutional Review Boards of Northwestern University (#STU00206507) and Midwestern University (#3047). The study took place at Northwestern Memorial Hospital (NMH), an 897-bed tertiary academic medical center in Chicago, IL. We conducted a retrospective, case-control study of patients treated empirically for CAP. Cases and controls were eligible if they were hospitalized between January 1, 2014 and May 29, 2018, were adults (aged 18 years or older), and received systemic antibiotics with an indication listed as CAP. Case patients were excluded if they were PCR positive for CDI within the previous 90 days, had cystic fibrosis, had a history of frequent hospitalization (≥3 admissions within prior 30 days), required initial ICU management for severe CAP, or had a rapidly fatal underlying condition (i.e., expired within 48 hours of admission). Inclusion and exclusion criteria for control patients were similar except that controls could not have had any prior documented positive *C. difficile* PCR test during the study period. The index admission for cases and controls was taken as the first hospitalization for CAP during the study period. All eligible cases were considered. Eligible controls were sampled from the entire cohort of CAP patients targeting a 7:1 ratio of controls for each case to maximize available power.

### C. difficile specimen processing and PCR methods

As previously described (16), *C. difficile* specimens at our facility were evaluated using PCR testing. Briefly, liquid stool specimens were screened and processed by the clinical microbiology laboratory. All formed stools were rejected by the laboratory. Specimens sent from patients with a previous PCR test result within the prior seven days were not orderable through the electronic health record (EHR) at our hospital. Evaluable clinical specimens underwent DNA extraction and PCR amplification for the toxin B gene (i.e., *tcdB)*. PCR was performed using one of the following methods according to study year: BD GeneOhm™ kit (BD Diagnostics) and SmartCycler (years 2012-2015) or BD Max™ kit (BD Diagnostics) (years 2015-2018). All PCR conditions were performed according to manufacturer protocol.

### Data Sources and Definitions

Patient data were retrieved from the EHR system (Cerner, Millennium) by querying the Northwestern Electronic Data Warehouse (EDW). Briefly, admissions records were searched for hospital encounters matching the inclusion criteria. Hospital admission characteristics (i.e., length of stay, antibiotic treatment), patient demographics (i.e., age, weight, sex, comorbidities) were extracted from the EHR. CDI was classified as a positive *C. difficile* PCR test. PCR test results were extracted from a line list of LabID events reported to the National Healthcare Safety Network (NHSN) accessed through the NMH portal. These data were cross-referenced against the list of all patients who were empirically treated for CAP during the study period. Patients were considered to be cases if they had a positive PCR test result on file within 180 days after the index admission for CAP. Controls could not have any positive PCR test results during the entire study period (i.e. 90 days before to 180 days after CAP diagnosis). CDI onset was classified according to NHSN criteria as community onset (CO), CO-healthcare facility associated (CO-HCFA), and hospital onset (HO). For the purposes of this study, we considered healthcare-associated CDI to be the composite of HO and CO-HFCA. According to the NHSN definitions, HO CDIs were defined as LabID events that occurred between day 4 of the index hospitalization and discharge, and CO-HCFA CDIs were defined as LabID events within 28 days after discharge from the index admission. Comorbidities and other clinical demographics at or before the index hospitalization were verified by manual chart review and included heart failure, dialysis-dependent chronic kidney disease, MRSA colonization, antibiotic receipt within the previous 90 days, hospitalization for two or more days in the previous 90 days, asthma, COPD, diabetes mellitus, ambulatory status, receipt of chronic acid suppression therapy, history of tube feeding requirement, hematologic malignancy, and receipt of immunosuppression therapy. Patient acuity and severity were classified using a modified-APACHE II score (m-APACHE II) (17) and the Pneumonia Severity Index (PSI) (18) applied on hospital day 1. The presence of chronic comorbidities including immunocompromise, respiratory failure, hepatic failure or heart failure, were assessed as a component of the m-APACHE II score. Data were stored in REDCap (19). CAP antibiotics were classified based on spectrum of activity, class, and as concordant or discordant based upon our institutional CAP treatment algorithm (**Figure S1**) (20). Broad-spectrum Gram-negative antibiotics included cefepime, meropenem, and piperacillin-tazobactam; Broad-spectrum Gram-positive antibiotics included linezolid and vancomycin; and guideline-concordant antibiotic use was considered empiric antibiotic use that followed our hospital’s CAP treatment algorithm.

### Univariate statistical analyses

Descriptive statistics were calculated for all variables. Differences in demographics between cases and randomly sampled controls were compared at the univariate level. Continuous variables were evaluated using Student’s t-test or Wilcoxon rank sum test and categorical variables were evaluated using Chi-square or Fisher’s exact tests, as appropriate. All analyses were performed in R (21).

### Model Development and Validation

Model development was completed using the *ODA* package for R (22), which is an R-based interface for the Optimal Data Analysis (ODA) and Classification Tree Analysis (CTA) software programs developed by Yarnold and Soltysik (23). Briefly, baseline attributes were evaluated during the index admission and considered as predictors of CDI case status for use in propensity score development. CO, CO-HFCA, HO, and any healthcare-associated CDI were evaluated as dependent (i.e., class) variables. Independent variables (i.e., attributes) observed post-admission (e.g., severity of illness and antibiotic treatment) were considered as independent variables to predict the class variables. Missing data were considered missing at random if univariate ODA failed to associate missingness with class membership. ODA identified variables with maximum accuracy for case status, and these were used in the propensity score weighting (24, 25). Propensity score weights were developed using baseline characteristics to address threats to causal inference (24). Time-to-event weights based on length of stay data were also considered as a weight alone or in combination with the propensity score weight (26). Next, CTA was used to identify a family of models that predicted CDI status using post-admission attributes, weighted by the optimal (i.e., having greatest effect strength after adjusting for chance) propensity score for each respective outcome. The sensitivity (Sn), specificity (Sp), and positive and negative predictive values for each class were calculated for each model. Model performance was described in terms of the Effect Strength for Sensitivity (ESS) which assesses predictive accuracy corrected for chance: for a two-category class variable, ESS=100 x [(0.5 x (Sp+Sn))-0.5]/0.5. In addition, the distance statistic D (23) is the number of additional effects with equivalent ESS needed to obtain perfect classification of the sample (27). The final models for each class variable were selected among competing models based upon their weighted ESS or WESS (indicating that observations are weighted with respect to their specific propensity and/or observed time) and D statistics. The model(s) with the highest WESS and lowest D was (were) considered the most accurate and parsimonious for each respective outcome. Cross-generalizability to an independent random sample was estimated via one sample (“leave-one-out” or LOO) jackknife analysis (28). The *NOVOboot()* function in ODA was used to evaluate the reliability of the predictions using bootstrap resampling with replacement (n=25,000 resamples with 50% replacement) to estimate an exact discrete 95% confidence interval (CI) for ESS for the model and this was compared to the exact discrete 95% CI for ESS for chance (i.e., model predictions were randomly scrambled relative to the class variable) (29).

## Results

### Sample Demographics

Overall, 4216 eligible patients received empiric treatment for CAP at our center during the study period. A total of 32 eligible PCR confirmed CDI cases were identified, and 232 randomly selected controls were drawn from the overall population of patients admitted during the study period. Cases were comprised of HO CDI (6.4%, n=17), CO CDI (3%, n=8), and CO-HCFA CDI (2.7%, n=7). Correspondingly, 9% (n=24) patients experienced a healthcare-associated CDI after their index CAP admission. Some data elements were missing for n=13 subjects, but these were considered as missing at random because univariate ODA failed to identify any associations with the above class variables (all P values > 0.2).

Baseline demographic differences between cases and controls are shown in **Table 1**. Case and controls in the training data set were similar with respect to female sex (50 vs 51%, respectively) and age [mean (SD): 68.5 (17.6) vs 62.4 (17.8), respectively]. However, cases had a higher mean (SD) modified-APACHE II score [11.2 (4.22) vs 8.14 (4.39); P<0.001] and a higher PSI [105 (26.3) vs 77.1 (31.2)]. In general, cases were more likely to have a history of acid suppressant use (P<0.01), tube feeding requirements (P<0.01), hematologic malignancy (P=0.02), immunosuppression (P=0.01), renal insufficiency (P<0.01), multiple chronic comorbidities (P<0.01), MRSA colonization (P=0.01), antibiotic use within the previous 90 days (P<0.001), and a history of previous hospitalization > 48 hr in the previous 90 days (P<0.001). Cases were more likely to have been admitted from a nursing home or LTAC (P=0.01), whereas controls were more likely to have been admitted to the general medicine service compared to oncology or surgical services (P<0.001).

**Table 1.** Demographics and baseline characteristics of cases and controls

|  | **Total**<br>**(N=264)** | **Case**<br>**(N=32)** | **Control**<br>**(N=232)** | **P-value** |
| --- | --- | --- | --- | --- |
| **Age (years)** |  |  |  |  |
| Mean (SD) | 63.1 (17.9) | 68.5 (17.6) | 62.4 (17.8) | 0.0705 |
| **Weight (kg)** |  |  |  |  |
| Mean (SD) | 84.4 (27.5) | 84.8 (28.6) | 84.4 (27.4) | 0.945 |
| Missing | 11 (4.2%) | 0 (0%) | 11 (4.7%) |  |
| **Temperature (highest in first 24 hr of admission)** |  |  |  |  |
| Mean (SD) | 37.3 (3.75) | 38.8 (10.3) | 37.1 (1.19) | 0.377 |
| **Respiratory rate (breaths/min)** |  |  |  |  |
| Mean (SD) | 19.5 (3.54) | 19.0 (3.10) | 19.5 (3.60) | 0.35 |
| Missing | 1 (0.4%) | 0 (0%) | 1 (0.4%) |  |
| **Heart rate (beats/min)** |  |  |  |  |
| Mean (SD) | 93.1 (18.6) | 95.2 (17.8) | 92.8 (18.7) | 0.487 |
| **White cell count (10<sup>3</sup> cells/μL)** |  |  |  |  |
| Mean (SD) | 10.0 (5.32) | 9.31 (5.75) | 10.1 (5.26) | 0.439 |
| Missing | 2 (0.8%) | 0 (0%) | 2 (0.9%) |  |
| **Modified APACHE II** |  |  |  |  |
| Mean (SD) | 8.52 (4.48) | 11.2 (4.22) | 8.14 (4.39) | <0.001 |
| Missing | 2 (0.8%) | 0 (0%) | 2 (0.9%) |  |
| **Pneumonia Severity Index** |  |  |  |  |
| Mean (SD) | 80.5 (32.0) | 105 (26.3) | 77.1 (31.2) | <0.001 |
| **Female** |  |  |  |  |
| n (%) | 135 (51.1%) | 16 (50.0%) | 119 (51.3%) | >0.99 |
| **African American/Black** |  |  |  |  |
| n (%) | 62 (23.5%) | 4 (12.5%) | 58 (25.0%) | 0.18 |
| **Asian** |  |  |  |  |
| n (%) | 7 (2.7%) | 1 (3.1%) | 6 (2.6%) | 0.6 |
| **Caucasian/White** |  |  |  |  |
| n (%) | 133 (50.4%) | 21 (65.6%) | 112 (48.3%) | 0.0888 |
| **Hispanic/Latinx** |  |  |  |  |
| n (%) | 9 (3.4%) | 1 (3.1%) | 8 (3.4%) | >0.99 |
| **Native American/Alaska Native** |  |  |  |  |
| n (%) | 1 (0.4%) | 1 (3.1%) | 0 (0%) | 0.121 |
| **Other Race** |  |  |  |  |
| n (%) | 51 (19.3%) | 4 (12.5%) | 47 (20.3%) | 0.35 |
| **Acute kidney injury** |  |  |  |  |
| n (%) | 30 (11.4%) | 3 (9.4%) | 27 (11.6%) | >0.99 |
| **Nursing home or LTAC resident** |  |  |  |  |
| n (%) | 5 (1.9%) | 3 (9.4%) | 2 (0.9%) | 0.0138 |

|  | Total<br>(N=264) | Case<br>(N=32) | Control<br>(N=232) | P-value |
| --- | --- | --- | --- | --- |
| **Altered mental status** |  |  |  |  |
| n (%) | 19 (7.2%) | 5 (15.6%) | 14 (6.0%) | 0.0637 |
| **Pleural effusion** |  |  |  |  |
| n (%) | 70 (26.5%) | 9 (28.1%) | 61 (26.3%) | 0.832 |
| **PORT Class II** |  |  |  |  |
| n (%) | 109 (41.3%) | 3 (9.4%) | 106 (45.7%) | <0.001 |
| **PORT Class III** |  |  |  |  |
| n (%) | 49 (18.6%) | 6 (18.8%) | 43 (18.5%) | >0.99 |
| **PORT Class IV** |  |  |  |  |
| n (%) | 89 (33.7%) | 17 (53.1%) | 72 (31.0%) | 0.0168 |
| **PORT Class V** |  |  |  |  |
| Yes | 17 (6.4%) | 6 (18.8%) | 11 (4.7%) | 0.00929 |
| **History of acid-suppressant use** |  |  |  |  |
| n (%) | 92 (34.8%) | 18 (56.2%) | 74 (31.9%) | 0.00957 |
| **History of asthma** |  |  |  |  |
| n (%) | 30 (11.4%) | 3 (9.4%) | 27 (11.6%) | >0.99 |
| **History of congestive heart failure** |  |  |  |  |
| n (%) | 24 (9.1%) | 6 (18.8%) | 18 (7.8%) | 0.0532 |
| **History of COPD** |  |  |  |  |
| n (%) | 39 (14.8%) | 4 (12.5%) | 35 (15.1%) | >0.99 |
| **History of Diabetes mellitus** |  |  |  |  |
| n (%) | 59 (22.3%) | 9 (28.1%) | 50 (21.6%) | 0.376 |
| **History of hematologic malignancy** |  |  |  |  |
| n (%) | 55 (20.8%) | 12 (37.5%) | 43 (18.5%) | 0.0195 |
| **History of immunosuppression<sup>a</sup>** |  |  |  |  |
| n (%) | 57 (21.6%) | 13 (40.6%) | 44 (19.0%) | 0.0101 |
| **History of renal insufficiency** |  |  |  |  |
| n (%) | 32 (12.1%) | 9 (28.1%) | 23 (9.9%) | 0.00708 |
| **History of tube-feeding requirement** |  |  |  |  |
| n (%) | 7 (2.7%) | 4 (12.5%) | 3 (1.3%) | 0.00486 |
| **Antibiotic use in the last 90 days** |  |  |  |  |
| n (%) | 83 (31.4%) | 19 (59.4%) | 64 (27.6%) | <0.001 |
| **MRSA colonization history** |  |  |  |  |
| n (%) | 2 (0.8%) | 2 (6.2%) | 0 (0%) | 0.0143 |
| **Non-ambulatory status** |  |  |  |  |
| n (%) | 6 (2.3%) | 0 (0%) | 6 (2.6%) | >0.99 |
| **Hospitalization > 48 hr in last 90 days** |  |  |  |  |
| n (%) | 56 (21.2%) | 18 (56.2%) | 38 (16.4%) | <0.001 |
| **Chronic comorbidities<sup>b</sup>** |  |  |  |  |
| n (%) | 74 (28.0%) | 17 (53.1%) | 57 (24.6%) | 0.00147 |
| Missing | 1 (0.4%) | 0 (0%) | 1 (0.4%) |  |
| **BUN > 29 mg/dL** |  |  |  |  |
| n (%) | 35 (13.3%) | 12 (37.5%) | 23 (9.9%) | <0.001 |
| **General medicine service** |  |  |  |  |
| n (%) | 252 (95.5%) | 23 (71.9%) | 229 (98.7%) | <0.001 |

**Abbreviations:** BUN, blood urea nitrogen; COPD, chronic obstructive pulmonary disease; LTAC, long-term acute care; MRSA, methicillin-resistant *Staphylococcus aureus*
**Legend:** a.) Chronic comorbidities included dialysis-dependent renal failure, advanced liver disease, symptomatic congestive heart failure, chronic respirator dependency, or immunocompromised status; b.) immunocompromised host defined by receipt of therapeutic immune suppression, chemotherapy, long-term or recent high dose corticosteroids, leukemia, lymphoma, or AIDS.

### Univariate Predictors of Case and Control Status

We evaluated a total of 39 baseline attributes as predictors of case or control classification, as shown in **Table 1**. ODA revealed seven attributes as having >24% ESS that also had stable estimates in jackknife (i.e., LOO) analysis. A summary of the attributes is shown in **Table 2**. Prior hospitalization > 48 hr in last 90 days (ESS=39.9%), modified APACHE II > 8.5 (ESS=32.9%), antibiotic use in the last 90 days (ESS=31.8%), the presence of chronic comorbidities (ESS=28.5%), BUN > 29 mg/dL (ESS=27.6%), and a history of acid-suppressant use (ESS=24.4%) were positively associated with case status whereas admission directly to a general medicine ward (ESS=26.8%) was associated with control status. These predictors were evaluated as potential propensity scores using classification tree analysis (CTA) to identify optimal weighted CTA models of CDI.

**Table 2.** Baseline attributes that were stable predictors of case or control in training and validity analysis

| Attribute, if “yes” then predict “Class” | Primary ODA Model |  |  |  | Leave-one-out Model |  |  |
| --- | --- | --- | --- | --- | --- | --- | --- |
|  | Class | PAC<br>% | ESS<br>% | M.C.<br>P-value | Class | ESS<br>% | M.C.<br>P-value |
| Hospitalization > 48 hr in last 90 days | Case | 80.3 | 39.9% | <0.001 | Case | 39.9% | 0.000003 |
| Modified-APACHE II > 8.5 | Case | 57.6 | 32.9% | 0.0016 | Case | 32.9% | 0.000382 |
| Antibiotic use in the last 90 days | Case | 70.8 | 31.8% | 0.0004 | Case | 31.8% | 0.000466 |
| Chronic comorbidities | Case | 72.6 | 28.5% | 0.0019 | Case | 28.5% | 0.001287 |
| BUN > 29 (mg/dL) | Case | 83.7 | 27.6% | 0.0002 | Case | 27.6% | 0.000165 |
| General medicine service | Control | 90.2 | 26.8% | <0.001 | Control | 26.8% | 0.00000032 |
| History of Acid-suppressant use | Case | 66.7 | 24.4% | 0.0103 | Case | 24.4% | 0.006898 |

**Legend:** M.C. P-values require no distributional assumptions and are exact.
**Abbreviations:** BUN, blood urea nitrogen; ESS, effect strength for sensitivity; ESP, effect strength for predictive value; M.C., Monte Carlo; PAC, percent accuracy in classification (i.e., overall accuracy).

Differences between cases and controls with respect to empiric antibiotic use are shown in **Table 3**. Guideline concordant antibiotic use was numerically more common among controls as compared to cases. Initial broad-spectrum Gram-negative agents were more commonly prescribed for cases than controls [n=11 (34.4%) vs n=24 (10.3%); P<0.001] as were broad-spectrum Gram-positive agents [n=11 (34.4%) vs n=25 (10.8%); P=0.00113].

**Table 3.** Receipt of broad-spectrum and guideline-concordant antibiotics among cases and controls

|  | Total<br>(N=264) | Case<br>(N=32) | Control<br>(N=232) | P-value |
| --- | --- | --- | --- | --- |
| Any broad-spectrum Gram-negative antibiotics |  |  |  |  |
| Yes | 35 (13.3%) | 11 (34.4%) | 24 (10.3%) | <0.001 |
| No | 229 (86.7%) | 21 (65.6%) | 208 (89.7%) |  |
| Any broad-spectrum Gram-positive antibiotics |  |  |  |  |
| Yes | 36 (13.6%) | 11 (34.4%) | 25 (10.8%) | 0.001 |
| No | 228 (86.4%) | 21 (65.6%) | 207 (89.2%) |  |
| Receipt of guideline-concordant antibiotics |  |  |  |  |
| Yes | 187 (70.8%) | 18 (56.2%) | 169 (72.8%) | 0.058 |
| No | 74 (28.0%) | 14 (43.8%) | 60 (25.9%) |  |
| Missing | 3 (1.1%) | 0 (0%) | 3 (1.3%) |  |

### Optimal propensity scores weights defined using Classification Tree Analysis

Of the baseline attributes identified as predictors of any CDI case status (n=32) in the univariate analysis, only prior hospitalization > 48 hr in the last 90 days and BUN > 29 mg/dL were retained after the first propensity score development step. Two potential configurations of these attributes were identified yielding two potential propensity scores: 1) a three strata CTA model rooted by prior hospitalization (ESS=53.12%), and 2) a three strata CTA model rooted by BUN > 29 mg/dL (ESS=53.12%). As these models were isometric with respect to predictive performance but differed in configuration, each was considered as a candidate propensity score in subsequent model building procedures. **Figure 1** shows the configuration and predictive performance of model (1) above. In addition to these two scores, a propensity score based on hospital length of stay (LOS), and the multiplicative interactions between LOS and each of the propensity scores derived from the two CTA models above were also considered (n=5 scores total). In this way, we identified every single statistically viable propensity score for the sample.

**Figure 1.**
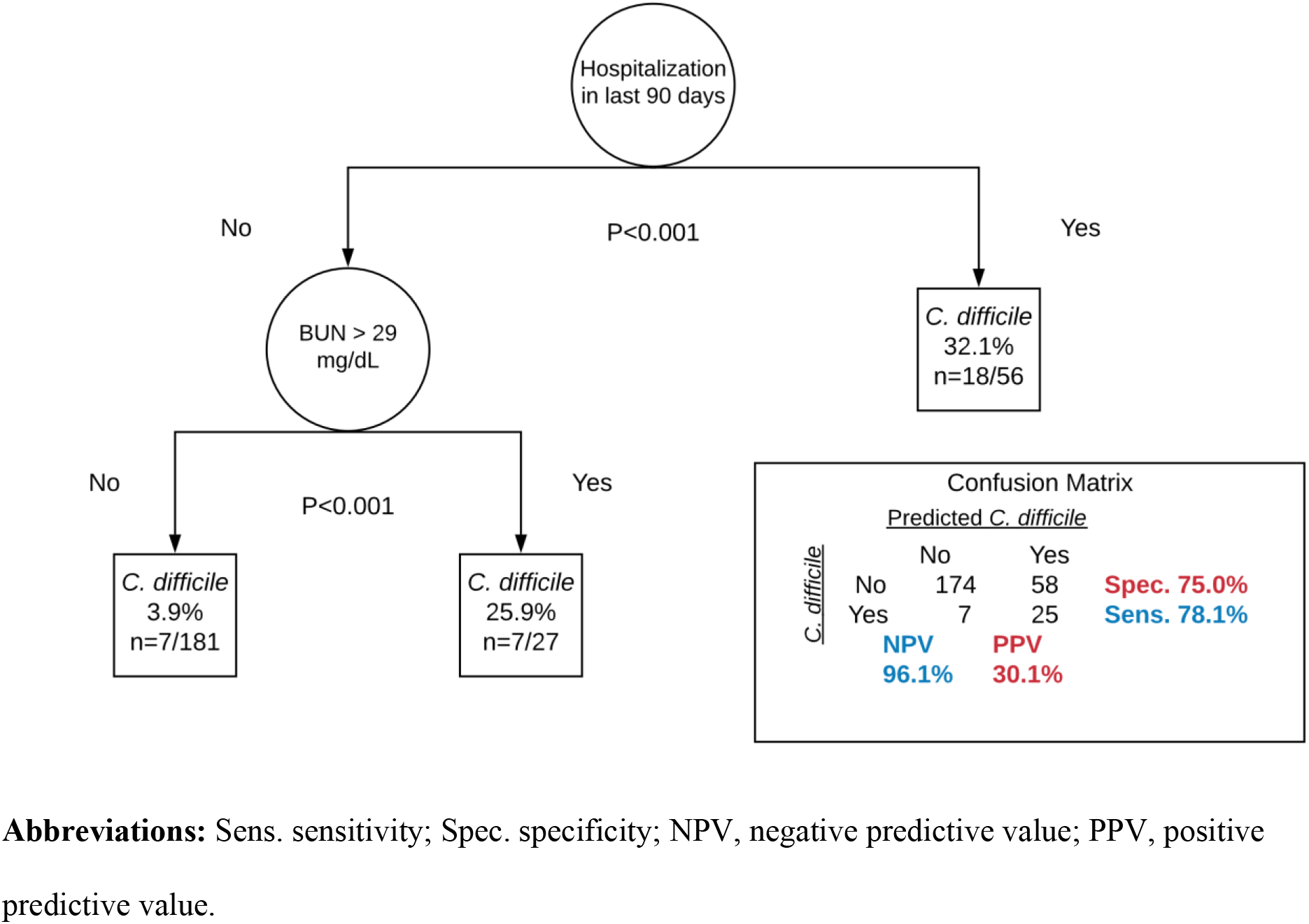
CTA propensity score model and associated model performance characteristics (Model 1)

### Selection of optimal Classification Tree Analysis Models for CDI

The results of the final model build are summarized in **Table 4**. Each of the following classifications were considered in model development: CO CDI, HO CDI, CO-HCFA CDI, and healthcare-associated CDI. The following attributes were considered as predictors in model development: empiric broad-spectrum Gram-negative antibiotic use, empiric broad-spectrum Gram-positive antibiotic use, guideline-concordant antibiotic use, and m-APACHE II score on the day of hospital admission. Each of the propensity scores developed above were iteratively considered and a weighted ESS (WESS) was calculated for each of the resulting models (n=5 propensity scores x n=4 classifications = up to 20 models to be evaluated), where the highest WESS maximizes overall model accuracy.

**Table 4.**
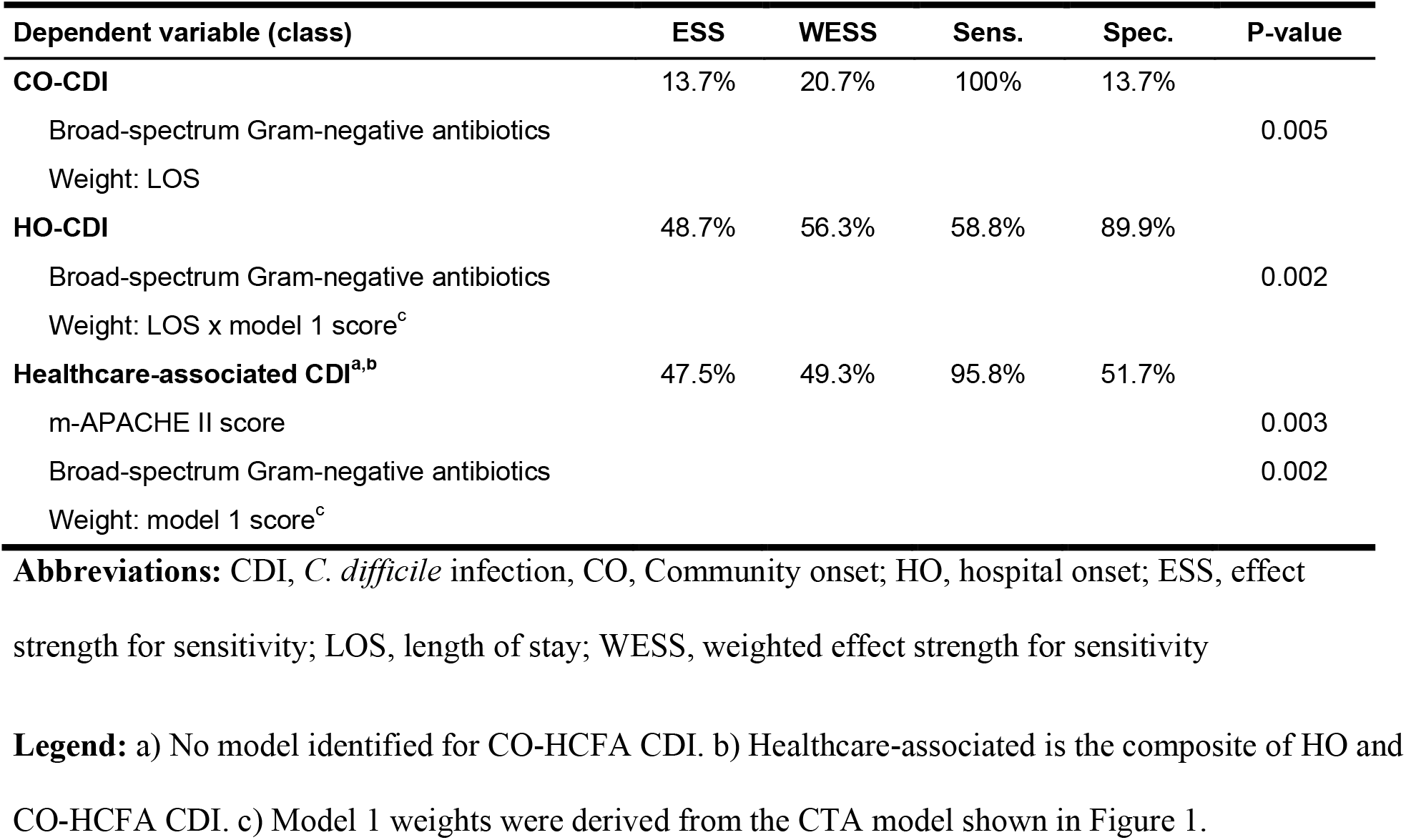
Summaries of the optimal weighted CTA models and performance metrics for each class

CTA identified only one predictive model for CO CDI; the model was weighted by LOS and had relatively low discriminatory power (ESS=13.7%, D=12.6, WESS=20.7%). CTA did not identify any predictive model for CO-HCFA CDI as a unique class. However, CTA identified several models for HO CDI. The globally optimal model predicted HO CDI using empiric broad-spectrum Gram-negative antibiotic use with moderate to strong discriminatory power (ESS=48.7%, D=2.1, WESS=56.3%). Likewise, CTA identified several models for healthcare-associated CDI; the globally optimal model predicted healthcare-associated CDI using m-APACHE II on hospital day 1 and empiric broad-spectrum Gram-negative antibiotic use with moderate discriminatory power (ESS=47.5%, D=3.3, WESS=49.3%).

The configuration and performance metrics of this model are shown in **Figure 2**. Each of these models had weighted PPV (wPPV) that exceeded the base rate of CDI for each class by over 2-fold (HO CDI base rate=6.4%, wPPV = 43.7%; CO CDI base rate=3%, wPPV=8.5%; and healthcare-associated CDI base rate=9%, wPPV=18.1%).

**Figure 2.**
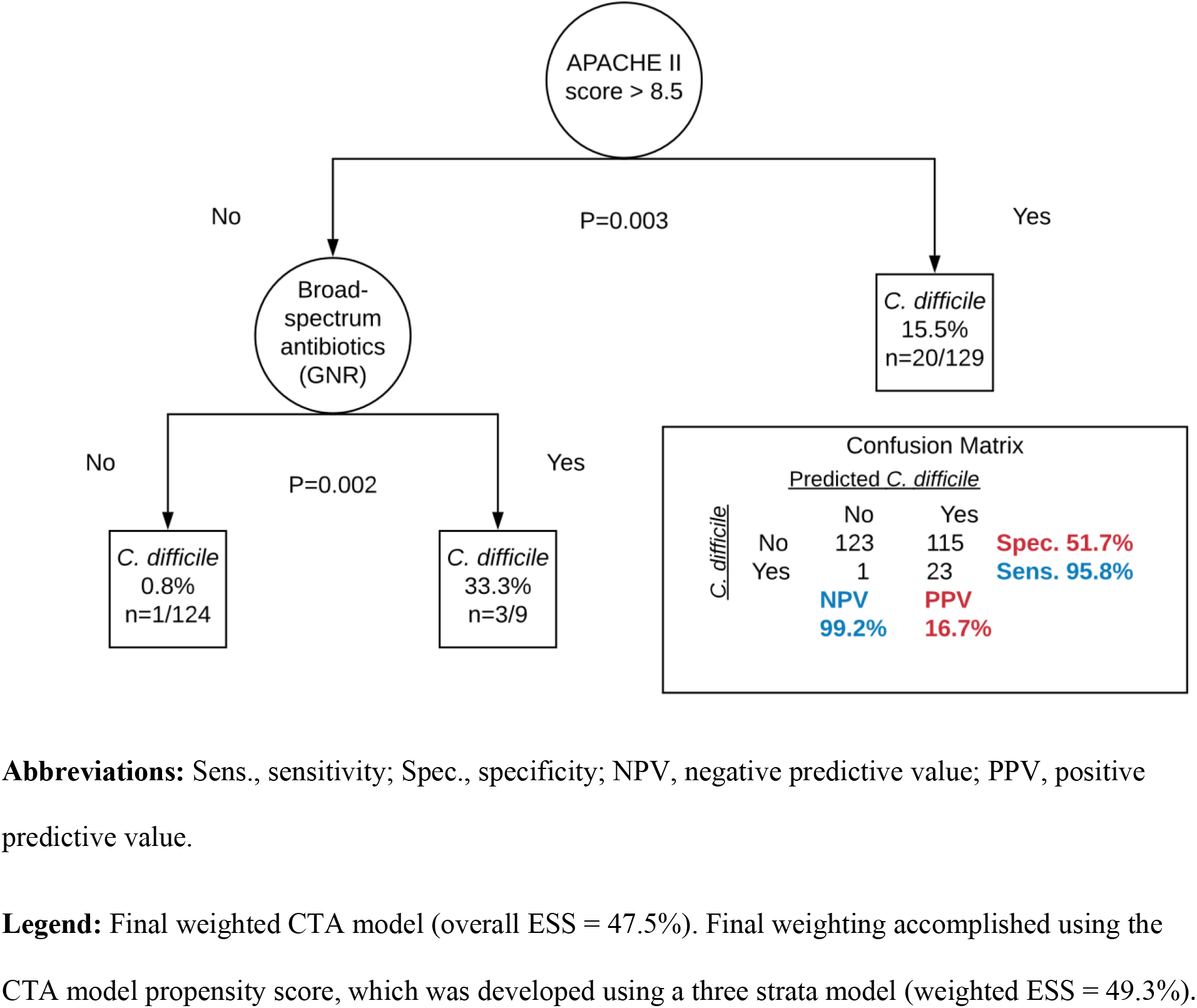
Globally optimal CTA model of healthcare-associated CDI and model performance characteristics (Final model)

### Validity assessment of the optimal Classification Tree Analysis Model

Novometric bootstrap validity assessment with n=25,000 resamples with 50% replacement was performed. In this analysis, the lower bound (2.5^th^ percentile) of the 95% CI for the model ESS and the upper bound (97.5^th^ percentile) of the 95% CI for chance ESS overlap was assessed. With respect to the models from **Table 4**, CO and HO CDI had model and chance 95% CI intervals that overlapped. No model was identified for CO-HCFA CDI. The model for healthcare-associated CDI had a lower bound 95% CI that did not overlap chance, confirming its generalizability. The results of this analysis are graphically depicted in **Figure 3**. These findings were reproduced in multiple random seeds (data not shown). Using this model, the odds of being a healthcare-associated CDI case were as follows: among those with an m-APACHE II ≤ 8.5 who did not receive broad-spectrum Gram-negative antibiotics, the odds were 1:125; among those with an m-APACHE II ≤ 8.5 who received broad-spectrum Gram-negative antibiotics the odds were 1:2, and among those with a m-APACHE II > 8.5 the odds were 5:27 independent of antibiotics prescribed for CAP.

**Figure 3.**
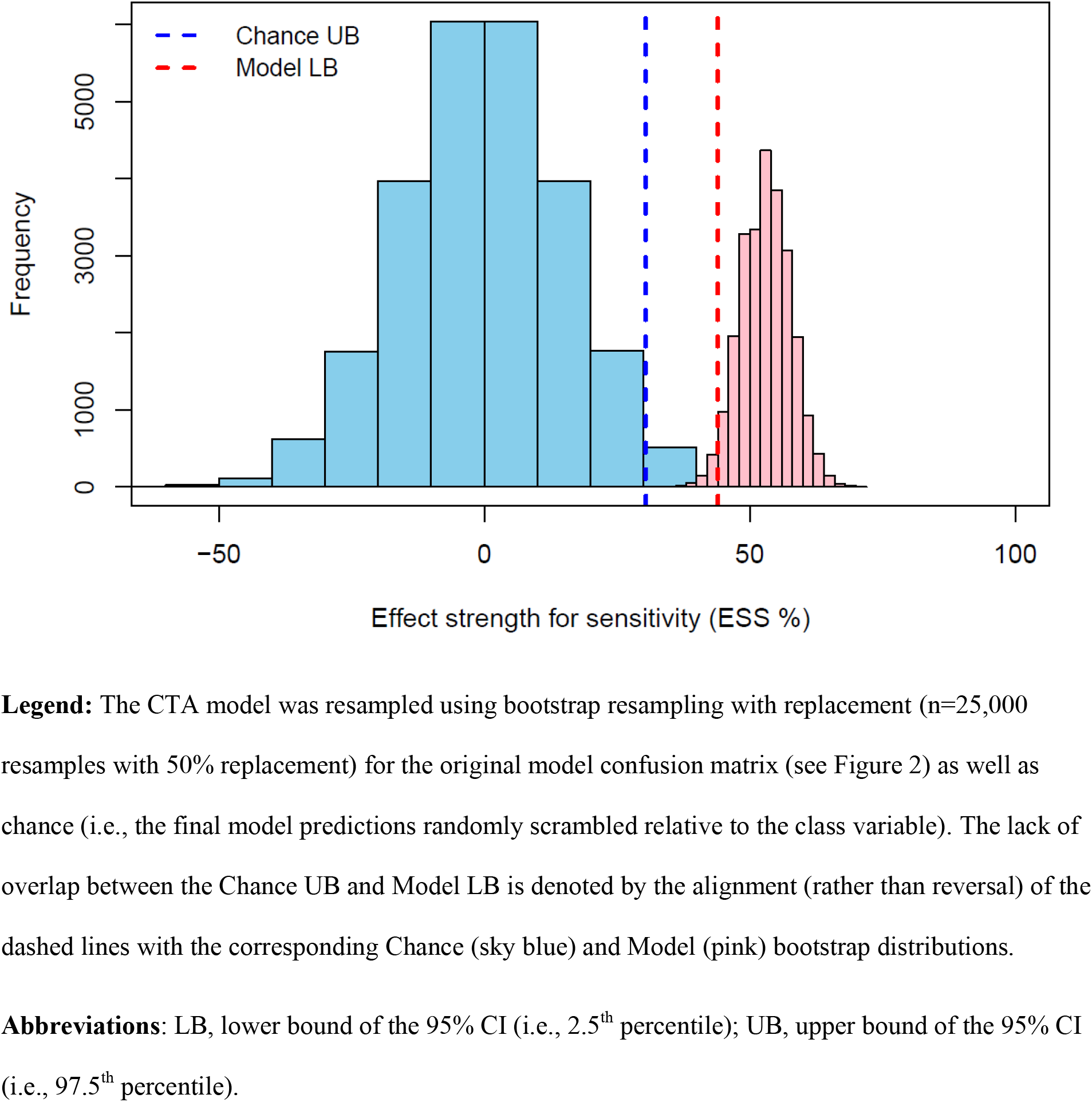
Novometric bootstrap analysis using the CTA model of healthcare-associated CDI (Final model).

## Discussion

We identified a series of predictive models that accounted for demographic, severity of illness, and post-admission factors that modify the risk of CDI in patients hospitalized for CAP. Broad-spectrum Gram-negative antibiotic use, including piperacillin-tazobactam, meropenem, and cefepime, was significantly associated with subsequent development of CO and HO CDI. Moreover, we identified a robust model of healthcare-associated (i.e., HO or CO-HCFA) CDI with high negative predictive value. Our models were weighted by optimal propensity scores for each outcome, and the PPV for each CDI model was more than 2-fold greater than the baseline CDI rate in each category. Our model of healthcare-associated CDI was reproducible and has implications for diagnostic *C difficile* PCR stewardship. For example, consider a patient who develops diarrhea while admitted to a hospital who 1) received only narrow-spectrum CAP antibiotics and 2) was not severely ill upon admission. In such a patient, a clinical decision rule could bypass the need for PCR-based testing given the low baseline risk of CDI, whereas a non-severely ill patient who received broad-spectrum antibiotics for CAP may benefit from PCR testing to rule out CDI. Our model also has the potential to impact empiric CAP antibiotic decision-making. For example, a 64-year-old male nursing home patient with past medical history of CHF who presented with a new cough, fever, and a focal infiltrate is admitted to the general floor for CAP. He is without confusion, tachypnea or history of liver disease or cancer. Electronic decision support notifies the prescriber that the patient’s m-APACHE II score is low and thus broad Gram-negative coverage for CAP dramatically increases the risk of developing CDI within the next few weeks for this patient. With this and other clinical and laboratory factors in mind, the prescriber may choose to initiate standard CAP antibiotic therapy and observe closely. Clearly, a critical appraisal of patient-specific factors is needed to fine-tune any diagnostic and prescribing algorithms.

Our findings have significance in the light of the revised pneumonia guidelines (30, 31). Healthcare-associated pneumonia (HCAP), a major driver of broad-spectrum antibiotic use, is no longer recognized as a valid diagnosis (31). The rationale for the abolition of HCAP stems from the poor predictive performance of these criteria when antibiotic-resistant pathogens are not endemic (32) and the failure of broad-spectrum agents to significantly improve outcomes among patients classified as having HCAP (14). Likewise, overuse of broad-spectrum antibiotics targeting HCAP can increase the risk of adverse effects such as CDI. We found that among patients with a lower m-APACHE II score who were empirically treated for CAP, receipt of broad-spectrum Gram-negative antibiotics increased the odds of healthcare-associated CDI to 1:2 compared to similar patients who did not receive these antibiotics (odds 1:125). Our predictive model is supported by multiple validity assessments including jackknife and bootstrap analyses. Our findings are consistent with prior research (33) and are biologically plausible. Patients treated for CAP experienced increased *C. difficile* carriage over the course of hospitalization, and the risk of carriage increases with excessive antibiotic treatment durations (10), which is supported by observational data (34). Broad-spectrum antibiotic use can also increase the risk of CDI. Chalmers et al. found that CDI was significantly associated with empiric use of piperacillin-tazobactam (22.9% vs 2.3%; P <0.0001) in patients with pneumonia. On the other hand, they found no significant association with CDI among patients receiving narrow-spectrum penicillins (24.6% vs 35.0%; P=0.09) (8). These findings are in line with ours and support the need for optimal approaches to antimicrobial stewardship for patients with CAP.

Our approach considered baseline factors including comorbidities, prior healthcare exposure, and baseline laboratory abnormalities as predictors of CDI. Our final propensity score consisted of hospitalization for > 48 hr in the previous 90 days at the root, with a secondary split for patients not previously hospitalized with an admission BUN > 29 mg/dL. We observed many differences between cases and controls in our univariate analysis. However, the majority of these factors 1) were unstable predictors in jackknife analysis or 2) were relatively weak effects. We considered baseline severity of illness (as measured by m-APACHE II score) as an adjustment factor in our final decision algorithms after propensity score weighting. Ultimately, m-APACHE II > 8.5 and broad-spectrum Gram-negative antibiotic use were major predictors of healthcare-associated CDI. HO CDI, specifically, was strongly associated with broad-spectrum Gram-negative antibiotic use. Thus, interventions that reduce broad-spectrum Gram-negative antibiotic use among patients being treated for CAP should reduce HO CDI.

Our study has several limitations. This was a single center retrospective case-control analysis. Cases are likely under-reported, and causality cannot be inferred. However, we identified suitable propensity score weights to address threats to causal inference and more appropriately balance cases and controls. Our final decision models are likely incomplete as evidenced by the D statistic (e.g., HO CDI: D=2.2, healthcare-associated CDI: D=3.3). Thus, two to three additional variables with equal explanatory power may exist that comprehensively classify each of these outcomes. More work is needed to improve clinical decision support related to *C. difficile* testing in this population. However, our decision model of healthcare-associated CDI risk was validated using several approaches, and this model can be easily translated into EMR-based algorithms and prescriber feedback strategies. Such an approach is supported by the current CDI guidelines, which call for institutional testing criteria to address false positives (3).

In summary, we identified relatively simple and biologically plausible clinical decision models that predict the risk of healthcare-associated CDI among hospitalized patients with CAP. Our models had high NPVs and, if paired with PCR testing in two-step clinical algorithms, could greatly reduce false positive PCR test results and avoid unnecessary treatment in patients colonized with *C. difficile*.

## Data Availability

Data will be made available upon reasonable request.

## Acknowledgements

None

## Funding Information

This research received no specific grant from any funding agency in the public, commercial, or not-for-profit sectors. This study was completed as part of our normal work. REDCap is supported at FSM by the Northwestern University Clinical and Translational Science (NUCATS) Institute. Research reported in this publication was supported, in part, by the National Institutes of Health’s National Center for Advancing Translational Sciences, Grant Number UL1TR001422. The content is solely the responsibility of the authors and does not necessarily represent the official views of the National Institutes of Health.

## Conflict of Interest Statement

The authors have no actual or potential conflicts of interest in relation to the data presented in this study.

## Data availability

Data will be made publicly available upon publication and upon request for peer review.

## Appendix

**Figure S1.**
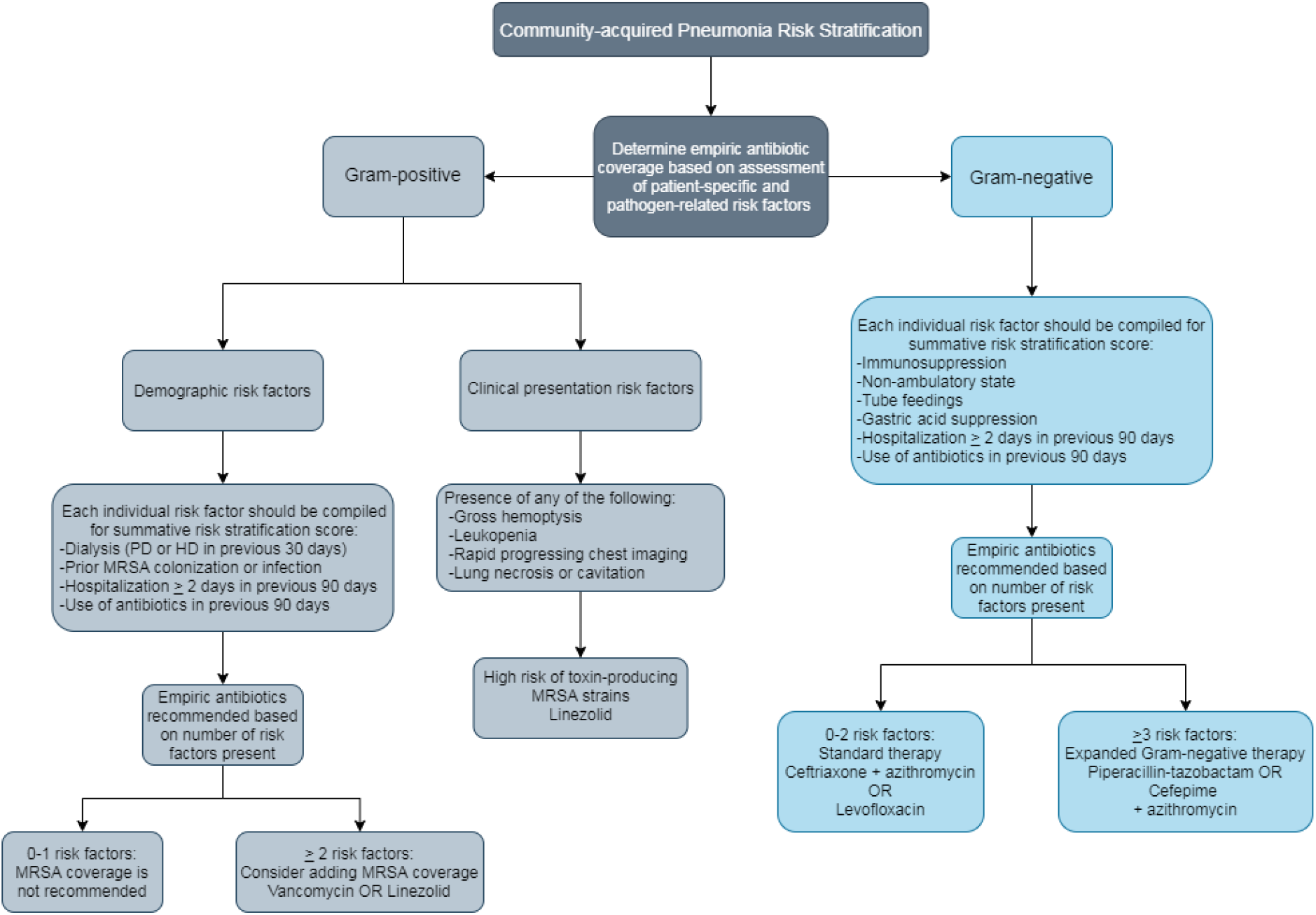
Northwestern Memorial Hospital CAP algorithm for non-ICU patients. Legend: Patients with two or fewer pre-defmed risk factors associated with respiratory infections caused by expanded spectrum Gram-negative pathogens *(Pseudomonas aeruginosa)* are recommended to receive standard empiric CAP therapy (e.g. ceftriaxone with azithromycin) and those patients with three or more risk factors associated with expanded spectrum Gram-negative pathogens are recommended to receive empiric broad-spectrum Gram-negative antibiotic coverage (e.g. piperacillin-tazobactam, cefepime) with a macrolide, as appropriate, based upon patient-specific drug allergies. Gram-positive risk necessitating empiric Gram-positive coverage (e.g. vancomycin, linezolid) is defined as the presence of two or more demographic risk factors or in those patients whose clinical presentation is consistent with MRSA pneumonia.

## Notes

### Competing Interest Statement

The authors have declared no competing interest.

### Author Declarations

Northwestern University IRB: protocol# STU00206507; Midwestern University IRB: protocol# 3017

